# Regional disparity in surgical residency training and work environment between urban and regional hospitals: an additional perspective from a nationwide survey for surgical residents

**DOI:** 10.1101/2024.12.11.24318834

**Authors:** Genki Watanabe, Jun Watanabe, Saki Hayashi, Takaaki Konishi, Saseem Poudel, Yoshiyuki Kiyasu, Masayuki Fukumoto, Takahiro Korai, Shinsuke Nomura, Naohiro Yamamoto, Masao Nakajima, Keiko Hosoya, Mitsue Saito

**Author notes:** Corresponding author: Jun Watanabe Department of Surgery, Division of Gastroenterological, General and Transplant Surgery, Jichi Medical University, Shimotsuke, Tochigi, Japan.

## Abstract

**Purpose:** To investigate the current status of regional disparity of surgical residency training between urban and regional hospitals.

**Methods:** Based on a nationwide online questionnaire survey for newly certified surgical trainees, the responding residents were divided into two groups according to the size of the city where they had trained: an urban city (>1 million, UC group) versus a regional city (<1 million, RC group). These groups were compared regarding surgical education and work environment.

**Results:** The UC group (n = 317, 42%) included surgeons with more post-graduate years, older age, females, and full-time working partners compared to the RC group (n = 439, 58%). More residents in the UC group were from urban areas, while the RC group had more residents from regional areas. No differences were observed in the number of experienced surgeries, published papers, opportunities for off-the-job training, or satisfaction with the surgical residency training program. Except for a higher income in the RC group, no differences were observed for other factors regarding the work environment.

**Conclusions:** There was little regional disparity regarding the impressions of surgical residency training between urban and regional cities including surgical education and work environment.

## Introduction

A surgeon shortage is a serious problem that makes surgical services inconvenient [1–3]. Japan is facing this dismal problem [4]. Indeed, the Japan Ministry of Health, Labour and Welfare (MHLW) indicated that the number of surgeons has fallen from 28,732 in 2000 to 27,634 in 2022 despite an increase in the total number of physicians [5]. This decline will have a pronounced impact on citizens in rural areas, where few surgeons work [6]. According to the list released by the Japan Surgical Society (JSS), the number of board-certified surgeons per 100,000 elderly people varied by prefectures from 35.4 to 100.1. Therefore, one of the keys to resolving the shortage is correcting surgeons’ disparity across regions [7]. This regional disparity of surgeons can occur during surgical residency after the initial two years of residency training following medical school graduation.

With the start of the “New Training and Certifying System of Medical Specialists in Japan”, the JSS revised the surgical residency training program in 2018, collaborating with the Japanese Medical Specialty Board [8, 9]. Under the new system, it is necessary to correct the regional disparity of surgeons. Jichi Medical University was established in 1972 for the purpose of ensuring doctors in rural areas and remote islands. The MHLW has expanded the regional quota for medical students who want to work in community medicine since 2008. Some local governments and private organizations have also established scholarship programs to secure future surgeons for working in regional areas. Despite these efforts, a nationwide survey conducted by the JSS to grasp the current distribution of surgeons in Japan reported that regional disparity remained a serious issue throughout Japan in 2021 [4].

The first nationwide needs assessment of surgical residency training in Japan was conducted by the Education Committee and Residency Curriculum Review Working Group of the JSS in 2016 [10, 11]. The additional analysis of this questionnaire survey identified regional disparity during surgical residency between training areas (large or small prefecture) and hospitals (university hospital or not) regarding training years, numbers of procedures, and competencies for procedures [12]. Although a revised surgical residency training program was launched and regional quotas and scholarship systems have been expanded, the current state of the regional disparity in surgical residency training is unclear.

The JSS educational committee U-40 working group conducted the second nationwide online survey about surgical residency training in 2023 [13]. As an additional analysis of the 2023 survey, the current study aimed to investigate whether regional disparity exists between urban and regional hospitals during surgical residency training including surgical education and work environment. The findings, highlighting disparity in surgical training between urban and regional hospitals, will help evaluate the outcomes of the 2018 revision of the surgical training program and may facilitate future solutions to the shortage of surgeons caused by regional disparity.

## Methods

### Surgical residency training for board-certified surgeons in Japan

After graduating from medical school and completing two years of the residency training program, residents aiming to become surgeons apply to the surgical training program at facilities certified by the JSS. During at least additional three years of the surgical training program, residents generally rotate through several hospitals in urban and rural areas. After completing the program, which includes a wide range of surgical experiences [over 350 surgeries, 120 as an operator] from gastrointestinal to trauma surgery, academic activities, conference presentations, and scholarly publications, residents take an examination to be a board-certified surgeon.

### Subjects of this study

This study reports findings from an additional analysis of the previously reported nationwide survey for all surgical residents who got certified in 2023. This research was approved by the institutional review board of the JSS (JSS2023-1). The online survey including 43 questions about the educational and work environment during surgical residency training was sent to the residents after their board-certification examination. All residents included in this study gave their informed consent to participate in this survey and were informed of their right to refuse their responses in advance.

The current analysis focuses on 4 specific questions associated with community medicine in the surgical field to clarify the actual impression and desirable factors for working at a regional hospital from the surgical residents’ perspective. One of the questions was an open-ended question asking about the desirable information and conditions for working in community medicine. In addition to these questions, the survey included questions about the residents’ hometown, training institution, and future workplace to clarify the current regional disparity during the residency training program. The details of the questions and results have been reported recently [13].

### Group definition to identify regional disparity in surgical residency training

The main training hospitals to which surgical residents belonged during the residency training program were divided into three groups according to the size of the city where the hospital is located based on Article 8 of the Local Autonomy Act [14]: the urban city group (UC, population over 1 million), the regional city group (RC, population from fifty thousand to 1 million), and the rural area group (RA, population within fifty thousand). Because residents who mainly worked at a hospital in RA were limited, we merged the RA group into the RC group. Therefore, this study finally compared the educational and work environment between the UC group and the RC group (including the RA group).

### Statistical analysis

In the tables, when the sum of the numbers for each variable does not match the UC group (n = 317) or the RC group (n = 439), this indicates that there are non-respondents. While categorical variables were compared using the χ^2^ test or Fisher’s exact test, continuous variables were shown as median (range) and were compared using the Mann–Whitney *U* test. Although questions regarding the satisfaction level were evaluated using a 4-level Likert scale (extremely satisfied, moderately satisfied, moderately dissatisfied, and extremely dissatisfied), the *p-*value was calculated on two levels: satisfied and dissatisfied. All tests were two-tailed, and *p* < 0.05 was considered statistically significant. Statistical analyses were conducted using IBM SPSS Statistics Version 28 (IBM Corporation, Armonk, NY, USA).

## Results

### Demographics of responding surgical residents

The response rate was 53.8% (758/1410). Among the 758 surgical residents, after 2 respondents were excluded due to insufficient answers, 756 eligible residents were divided into the UC group (n = 317, 41.9%) and the RC group (n = 439, 58.1%). The RC group included 29 (3.8%) surgical residents who underwent surgical training in RA. The flow of these residents from their hometowns to training institutions and future workplaces is illustrated in Figure 1.

**Fig. 1.**
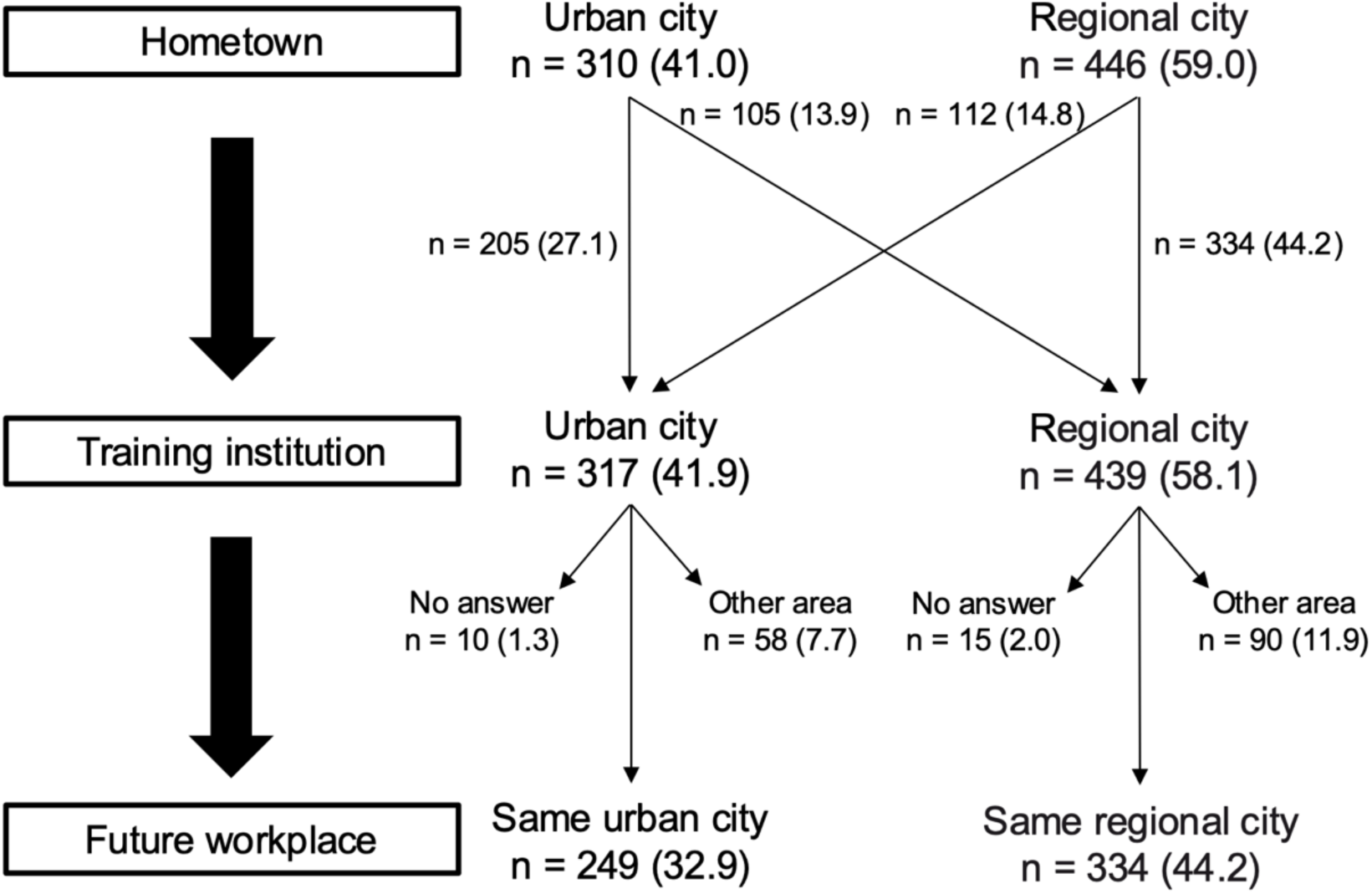
Flowchart showing the transfers of surgical residents from their hometowns to hospitals at surgical residency and eventually to their future workplaces. Numbers in parentheses indicate percentages derived from a total of 756 respondents.

Table 1 shows the demographics of the responding residents. All residents had obtained their medical licenses between 1989 and 2019. Surgeons who obtained medical licenses after 2016 comprised 83.8% in the UC group and 92.2% in the RC group (*p* < 0.001). The UC group included surgeons with longer post-graduate years, older age, and female sex compared with the RC group (all *p* < 0.001). Residents in the UC group were more likely to have a full-time working spouse or partner (*p* = 0.025). More residents (64.7%) in the UC group were from urban areas, while the RC group had more residents (62.6%) from regional areas (*p* < 0.001). The RC group had more residents who wanted to be gastroenterological surgeons than the UC group (59.9% vs. 42.3%, *p* < 0.001). Regional quotas or scholarships were selected as the most influential reason for deciding a surgical residency training program by 1.3% (4/317) of the UC group and 5.2% (23/439) of the RC group (Figure 2).

**Table 1.** Demographics of the responding surgical residents

| Variables | UC group<br>n = 317 | RC group<br>n = 439 | *p* |
| --- | --- | --- | --- |
| Post-graduation, years |  |  | <0.001 |
| 6 | 116 (36.6) | 199 (45.3) |  |
| 7 | 138 (43.5) | 195 (44.4) |  |
| Others | 63 (19.9) | 45 (10.3) |  |
| Age, years |  |  | <0.001 |
| ≤30 | 29 (9.1) | 88 (20.0) |  |
| 31–35 | 256 (80.8) | 315 (71.8) |  |
| 36–40 | 27 (8.5) | 30 (6.8) |  |
| >40 | 5 (1.6) | 6 (1.4) |  |
| Sex, female | 107 (33.8) | 87 (19.8) | <0.001 |
| Marital status married/with partner | 222 (70.0) | 319 (72.7) | 0.462 |
| Number of respondents with child(ren) | 124 (39.1) | 189 (43.1) | 0.310 |
| Full-time working spouse/partner | 151 (47.6) | 172 (39.2) | 0.025 |
| Belong to a department at a university hospital | 248 (78.2) | 355 (80.9) | 0.425 |
| Hometown |  |  | <0.001 |
| Urban city | 205 (64.7) | 105 (23.9) |  |
| Regional city | 90 (28.4) | 275 (62.6) |  |
| Rural area | 22 (6.9) | 59 (13.4) |  |
| Subspecialty of interest |  |  | <0.001 |
| Gastroenterological surgery | 134 (42.3) | 263 (59.9) |  |
| Cardiovascular surgery | 44 (13.9) | 42 (9.6) |  |
| Thoracic surgery | 44 (13.9) | 41 (9.3) |  |
| Breast surgery | 43 (13.6) | 42 (9.6) |  |
| Pediatric surgery | 24 (7.6) | 24 (5.5) |  |
| Acute care surgery | 16 (5.0) | 10 (2.3) |  |
| Endocrine surgery | 2 (0.6) | 6 (1.4) |  |
| Other | 10 (3.2) | 11 (2.5) |  |

Data are shown as n (%). UC, urban city; RC, regional city

**Fig. 2.**
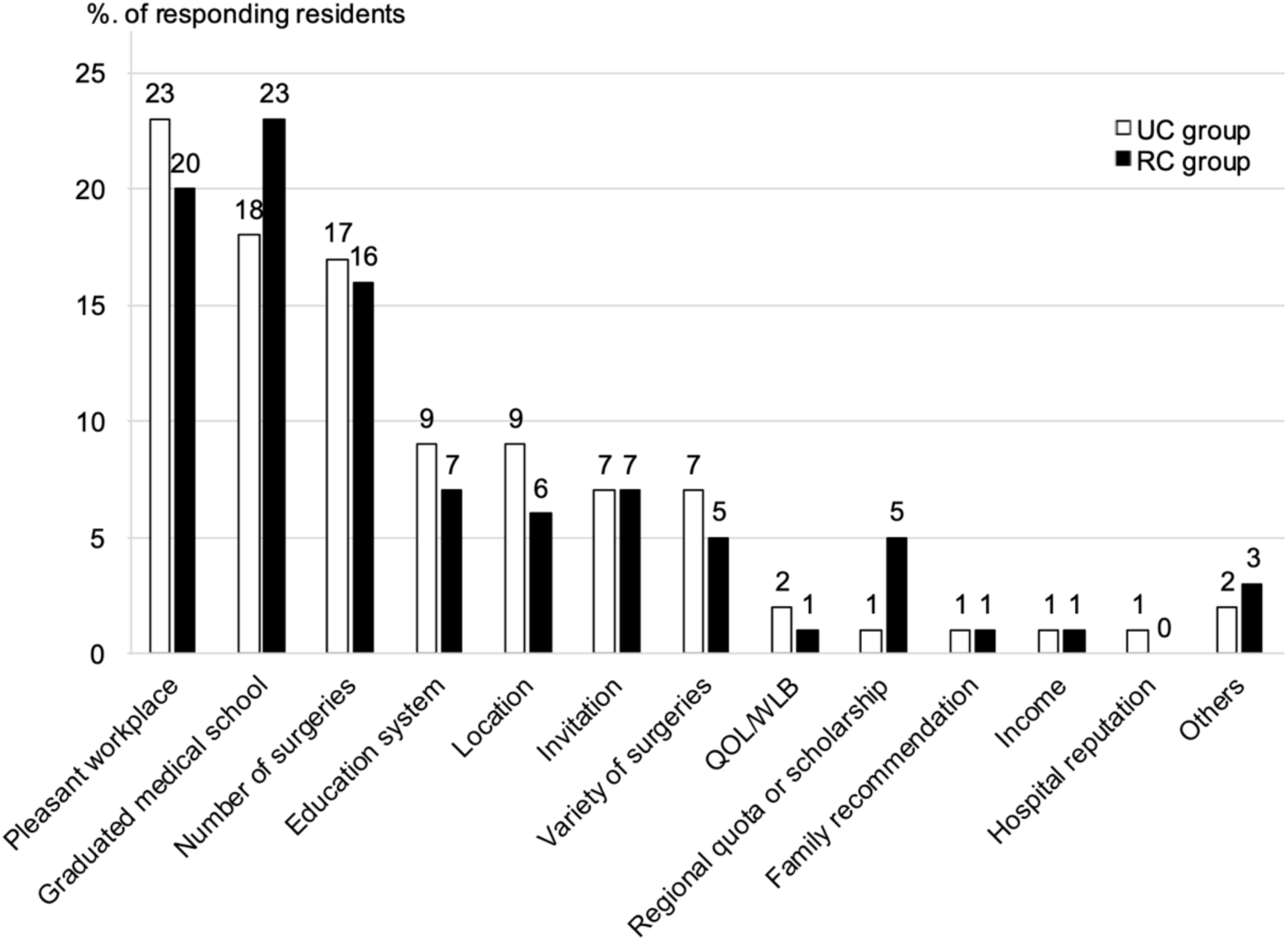
Most influential reasons for selecting the surgical residency program. Numbers above the bars indicate the percentage of responding residents. UC, urban city; RC, regional city; QOL, quality of life; WLB, work-life balance

### Educational environment during surgical residency training

Although there was no difference in belonging to a department at a university hospital during surgical residency training between the UC and the RC group (78.2% vs. 80.9%, *p* = 0.425), the period spent at an academic hospital such as a university hospital statistically differed between the two groups (*p* < 0.001). No statistical differences were observed in the period spent at tertiary or other hospitals, number of experienced surgeries under general anesthesia, areas difficult to experience, number of published papers, or opportunities for off-the-job training including technical or non-technical skills (Table 2).

**Table 2.** Comparison of surgical residency program between urban and regional hospitals

| Variables | UC group<br>n = 317 | RC group<br>n = 439 | *p* |
| --- | --- | --- | --- |
| Training institute/academic (months) |  |  | <0.001 |
| ≤6 | 45 (14.2) | 75 (17.1) |  |
| 7–12 | 79 (24.9) | 148 (33.7) |  |
| 13–24 | 45 (14.2) | 84 (19.1) |  |
| 25–36 | 37 (11.7) | 43 (9.8) |  |
| >36 | 37 (11.7) | 25 (5.7) |  |
| None | 59 (18.6) | 50 (11.4) |  |
| Training institute/tertiary (months) |  |  | 0.580 |
| ≤6 | 15 (4.7) | 18 (4.1) |  |
| 7–12 | 42 (13.2) | 79 (18.0) |  |
| 13–24 | 53 (16.7) | 63 (14.4) |  |
| 25–36 | 57 (18.0) | 79 (18.0) |  |
| >36 | 34 (10.7) | 42 (9.6) |  |
| None | 90 (28.4) | 131 (29.8) |  |
| Training institute/other (months) |  |  | 0.510 |
| ≤6 | 46 (14.5) | 55 (12.5) |  |
| 7–12 | 52 (16.4) | 60 (13.7) |  |
| 13–24 | 48 (15.1) | 85 (19.4) |  |
| 25–36 | 52 (16.4) | 86 (19.6) |  |
| >36 | 29 (9.1) | 45 (10.3) |  |
| None | 47 (14.8) | 68 (15.5) |  |
| Number of general anesthesia operations |  |  | 0.140 |
| ≤100 | 12 (3.8) | 16 (3.6) |  |
| 101–200 | 88 (27.8) | 90 (20.5) |  |
| 201–300 | 92 (29.0) | 132 (30.1) |  |
| 301–400 | 57 (18.0) | 107 (24.4) |  |
| 401–500 | 21 (6.6) | 35 (8.0) |  |
| ≥501 | 46 (14.5) | 58 (13.2) |  |
| Area difficult to experience |  |  | 0.220<sup>a</sup> |
| Trauma | 131 (41.3) | 160 (36.4) |  |
| Pediatric | 42 (13.2) | 49 (11.2) |  |
| Cardiovascular | 34 (10.7) | 56 (12.8) |  |
| Peripheral vascular | 23 (7.3) | 39 (8.9) |  |
| Head, neck, endocrine | 17 (5.4) | 24 (5.5) |  |
| Gastroenterology | 9 (2.8) | 10 (2.3) |  |
| Breast | 4 (1.3) | 5 (1.1) |  |
| Thoracic | 3 (0.9) | 14 (3.2) |  |
| Had no problem | 46 (14.5) | 66 (15.0) |  |
| Number of published papers |  |  | 0.500 |
| 0 | 71 (22.4) | 98 (22.3) |  |
| 1 | 101 (31.9) | 139 (31.7) |  |
| 2 | 50 (15.8) | 86 (19.6) |  |
| 3 | 35 (11.0) | 47 (10.7) |  |
| 4 | 18 (5.7) | 24 (5.5) |  |
| ≥5 | 33 (10.4) | 30 (6.8) |  |
| Off-the-job training/technical skills |  |  | 0.080 |
| Undertaken regularly | 66 (20.8) | 61 (13.9) |  |
| Undertaken inconsistently | 213 (67.2) | 309 (70.4) |  |
| Did not take the opportunity | 9 (2.8) | 9 (2.1) |  |
| Did not have the opportunity | 20 (6.3) | 41 (9.3) |  |
| Unsure | 2 (0.6) | 4 (0.9) |  |
| Off-the-job training/non-technical skills |  |  | 0.310 |
| Undertaken regularly | 28 (8.8) | 22 (5.0) |  |
| Undertaken inconsistently | 68 (21.5) | 101 (23.0) |  |
| Did not take the opportunity | 13 (4.1) | 14 (3.2) |  |
| Did not have the opportunity | 127 (40.1) | 183 (41.7) |  |
| Unsure | 74 (23.3) | 102 (23.2) |  |

Data are shown as n (%).
<sup>a</sup>*P*-values were calculated for the incidence of residents who had difficulties experiencing trauma cases.
UC, urban city; RC, regional city

In addition to the actual educational environment, subjective evaluation for the current surgical residency training program was comparable between the two groups. Namely, residents in both groups were similarly satisfied with the instructors’ clinical skills, educational skills, and surgical residency training program (*p* = 0.999, 0.915, and 0.999, respectively) (Figure 3).

**Fig. 3.**
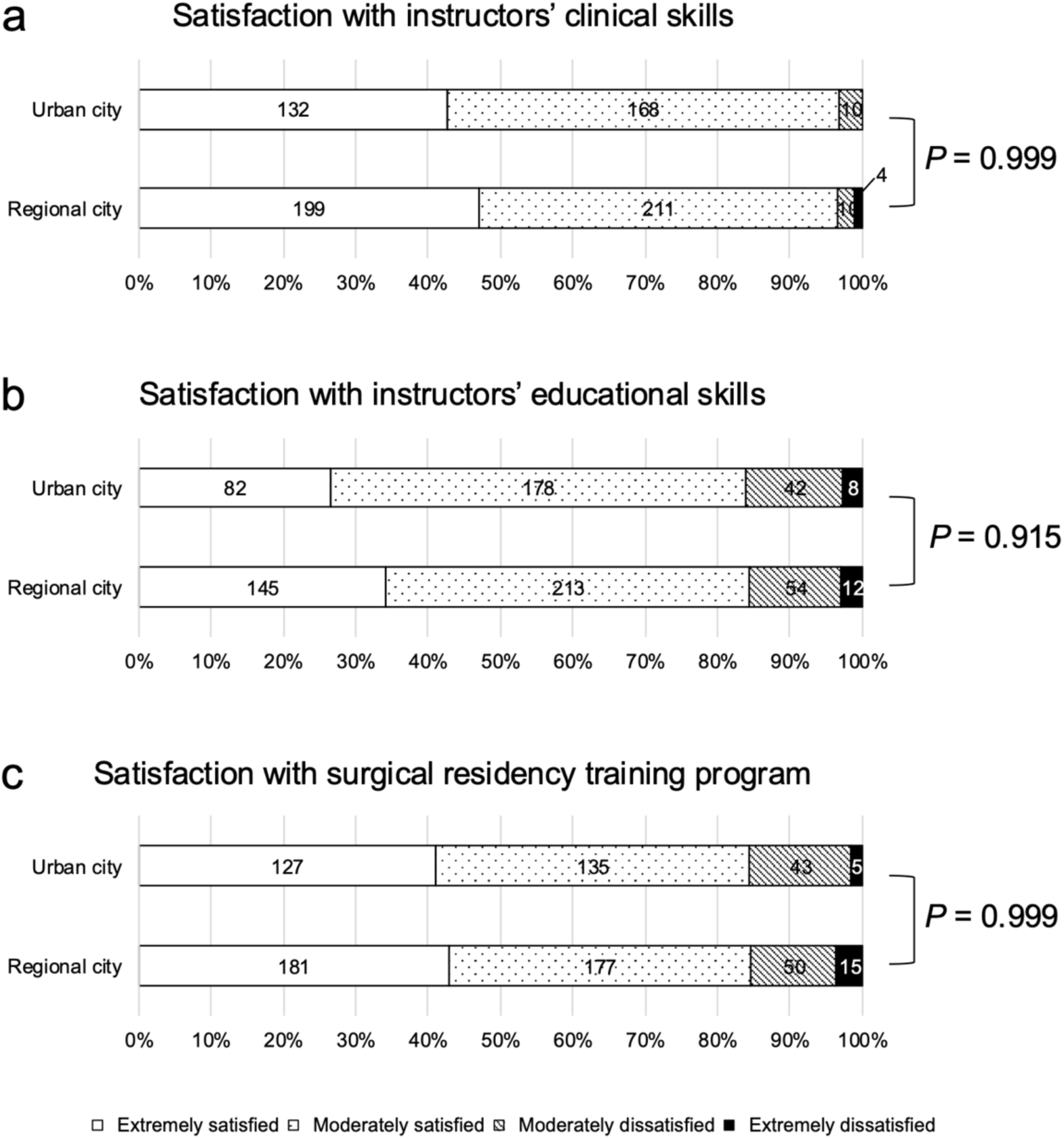
Satisfaction with instructors’ clinical skills (**a**), instructors’ educational skills **(b),** and surgical residency training program (**c**). Numbers inside the bars indicate the percentage of responding residents.

### Work environment during surgical residency training

Both groups had comparable overtime work per month with well-managed working hours (Table 3). The percentage of residents who believed that they had experienced some form of harassment did not differ, with 44.2% in the UC group and 37.1% in the RC group (*p* = 0.950). While night duty days (median) per month were comparable between the two groups (both 5 days, *p* = 0.050), the median annual income in the RC group was higher than that in the UC group (10 million yen vs. 9 million yen, *p* < 0.001).

**Table 3.** Comparison of work environment between urban and regional hospitals

| Variables | UC group<br>n = 317 | RC group<br>n = 439 | *p* |
| --- | --- | --- | --- |
| Overtime work per month, hours |  |  | 0.500 |
| ≤40 | 11 (3.5) | 16 (3.6) |  |
| 41–80 | 90 (28.4) | 114 (26.0) |  |
| 81–120 | 121 (38.2) | 167 (38.0) |  |
| 121–160 | 33 (10.4) | 64 (14.6) |  |
| ≥161 | 37 (11.7) | 40 (9.1) |  |
| Unknown | 12 (3.8) | 20 (4.6) |  |
| Working hour management |  |  | 0.080 |
| Strictly managed (time card, etc.) | 50 (15.8) | 52 (11.8) |  |
| Inadequately managed | 157 (49.5) | 254 (57.9) |  |
| Not managed | 91 (28.7) | 111 (25.3) |  |
| Do not know | 5 (1.6) | 3 (0.7) |  |
| Night duty days per months, days | 5 (0–31) | 5 (0–21) | 0.050 |
| Annual income (×1 million yen)<sup>a</sup> | 9 (1–38) | 10 (0–27) | <0.001 |
| Experienced any harassment during training |  |  | 0.950 |
| No | 154 (48.6) | 251 (57.2) |  |
| Yes | 140 (44.2) | 163 (37.1) |  |
| Do not want to answer | 23 (7.3) | 25 (5.7) |  |

Data are shown as n (%) or median (range).
<sup>a</sup>Annual income was calculated at the primary facility in the final year of the residency training.
UC, urban city; RC, regional city

### Desirable factors for working as surgeons at a regional hospital

More residents in the UC group had a negative impression of surgical residency training at regional hospitals than in the RC group (39.4% vs. 22.8%, *p* < 0.001) (Table 4). From the perspective of surgical residents, the factors affecting positive or negative impressions for surgical residency training at regional hospitals varied between the two groups. Although some differences between the two groups were observed, the top listed factors affecting positive impressions were common in both groups. They included income, number of surgeries performed, interpersonal relationships, high quality of life (QOL)/work life balance (WLB), and variety of surgeries. Similarly, the top listed factors affecting negative impressions were also common in both groups. They included educational system, few opportunities to perform treatment according to guidelines, workplace environment excluding interpersonal relationships, poor QOL/WLB, and impact on achievements and career (Table 4).

**Table 4.** Impression and details of surgical residency training at regional hospitals

| Variables | UC group<br>n = 317 | RC group<br>n = 439 | *p* |
| --- | --- | --- | --- |
| Impression of surgical residency training at regional hospitals |  |  | <0.001 |
| Positive impression | 192 (60.6) | 339 (77.2) |  |
| Negative impression or no answer | 125 (39.4) | 100 (22.8) |  |
| Factor of positive impression<sup>a</sup> |  |  |  |
| Income | 167 (52.7) | 196 (44.6) | 0.036 |
| Numbers of surgeries performed | 152 (47.9) | 253 (57.6) | 0.008 |
| Interpersonal relationships | 93 (29.3) | 157 (35.8) | 0.064 |
| QOL, WLB | 92 (29.0) | 117 (26.7) | 0.532 |
| Variety of surgeries | 91 (28.7) | 174 (39.6) | 0.002 |
| Workplace environment excluding interpersonal relationships | 36 (11.4) | 91 (20.7) | <0.001 |
| Educational system | 14 (4.4) | 38 (8.7) | 0.023 |
| Impact on achievements and career | 7 (2.2) | 11 (2.5) | 0.791 |
| Nothing | 8 (2.5) | 5 (1.1) | 0.148 |
| Factor of negative impression<sup>a</sup> |  |  | 0.02 |
| Educational system | 132 (41.6) | 148 (33.7) | 0.032 |
| Few opportunities to perform treatment according to guidelines | 101 (31.9) | 97 (22.1) | 0.003 |
| Workplace environment excluding interpersonal relationships | 96 (30.3) | 74 (16.9) | <0.001 |
| QOL, WLB | 95 (30.0) | 137 (31.2) | 0.716 |
| Impact on achievements and career | 86 (27.1) | 104 (23.7) | 0.282 |
| Numbers of surgeries performed | 86 (27.1) | 90 (20.5) | 0.033 |
| Variety of surgeries | 73 (23.0) | 99 (22.6) | 0.877 |
| Interpersonal relationships | 46 (14.5) | 45 (10.3) | 0.098 |
| Income | 10 (3.2) | 18 (4.1) | 0.497 |
| Nothing | 13 (4.1) | 32 (7.3) | 0.068 |

Data are shown as n (%).
<sup>a</sup>This was a multiple-choice question.
UC, urban city; RC, regional city; QOL, quality of life; WLB, work life balance

A total of 289 residents (38.1% of all 758 responding residents) responded to an open-ended question asking about desirable information and conditions for working at regional hospitals. The top five favorable conditions for community medicine were income (n = 87, 30.1%), QOL/WLB (n = 73, 25.3%), sufficient numbers of surgeries (n = 68, 23.5%), a well-organized education system (n = 62, 21.5%), followed by work environment excluding interpersonal relationships (n = 33, 11.4%) (Figure 4).

**Fig. 4.**
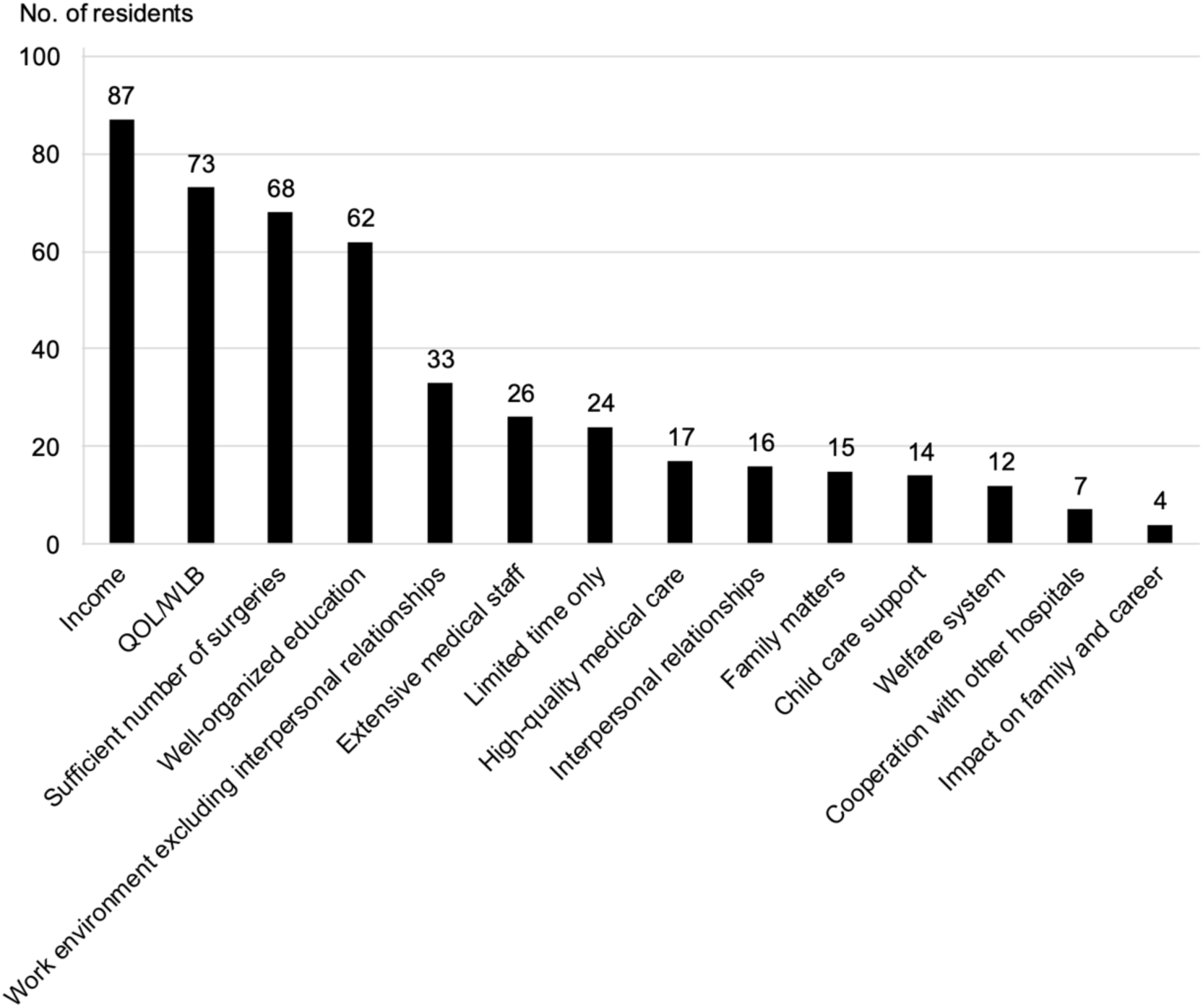
Desirable information and conditions for working at regional hospitals. Numbers above the bars indicate the actual number of responding residents. QOL, quality of life; WLB, work-life balance

## Discussion

This nationwide survey for surgical residents revealed the current status of regional disparity during surgical residency training in Japan. No statistical differences in surgical residency training were observed between urban and regional cities in terms of surgical experiences, academic works, night duty work, overtime work, or satisfaction with instructors and the surgical residency program. The annual income in the RC group was higher than that in the UC group. These results showed that there were no differences, except for a few items, for educational and work environment during surgical residency training between urban and regional cities.

Revisions made to the surgical residency training programs in 2018 may contribute to narrow the differences in surgical residency programs between urban and regional cities, which may help solve the surgeon shortage due to regional disparity. Previous reports identified several factors that encourage working in rural areas, such as a rural background, early exposure to rural practice, incentives, post-graduate education, and family matters [6, 15–17]. In our study, concordance between hometown and training institution were commonly observed in both the UC group (65%) and the RC group (76%).

Moreover, most residents in the UC group (79%) and the RC group (76%) chose their future workplace to be in the same area as the training institution. This trend was consistent with a previous paper reporting the effect of a physician’s hometown on the practice location [18]. Therefore, recruitment of surgical residents with regional or rural backgrounds could be an effective strategy for increasing the workforce in these areas. It has been reported that early exposure of medical students or residents to community medical care increases the possibility of working as a surgeon in regional areas in the future [19, 20]. Therefore, working in a regional city for a certain period under the revised surgical residency training may help to eliminate regional disparity by increasing the number of surgeons working there. The annual income in the RC group was higher than that in the UC group with a median difference of 1 million yen, despite similar levels of overtime work and night duty. However, it is unclear whether this difference is attributable to incentives for rural work. A more detailed survey is warranted in the future. Regarding regional quotas and scholarships, these were selected as reasons for choosing the training program in 5% of the RC group, and it is assumed that funding had a certain effect on securing surgeons to work in regional areas as previously reported [17]. Other encouraging factors for working in rural areas such as post-graduate education and family matters were not clarified in this study and are complex to deal with. Another finding of this study was a discrepancy between the impression and the actual status of working at regional hospitals, especially in the UC group. In other words, no differences were observed in the comparison of surgical residency training between urban and regional areas, although residents in the UC group had a statistically more negative impression of training at a regional hospital compared to those in the RC group regarding education system and work environment. Identifying the reasons for and resolving this discrepancy may provide an opportunity for surgeons trained in urban cities to direct their attention to working in regional areas, leading to solving the shortage in regional areas.

A previous paper identified the regional disparity between training areas (large or small prefecture) and hospitals (university hospital or not) in surgical resident training [12]. Our regional grouping (UC versus RC) did not follow the grouping used in this previous paper due to the differences in survey questions, which might require caution when comparing our study with previous studies. Any regional disparity in surgical resident training was not observed in the current study regarding number of surgeries experienced, difficulty to experience trauma cases, and opportunities to receive off-the-job training, which were opposite results compared to the previous report [12]. The reason for this discrepancy is not apparent, but it might be partially related to the revised surgical residency training programs which encourage having a variety of surgical experiences not only at a single institution but also at multiple institutions.

It is noteworthy that this survey directly inquired about surgical residents’ desirable information and conditions for working at regional hospitals. Modern surgical residents in Japan desire not only an appropriate surgical education and work environment, but also QOL and WLB which were ranked second as desirable factors to work at regional hospitals. Lifestyle factors were also the most common cause of attrition among surgical residents in other countries [21, 22]. Recently, achievement of WLB and QOL in surgeons raised concerns against the stereotypical surgeon lifestyle [23]. Because uncontrollable lifestyle factors were associated with burnout rates in surgeons [24], efforts to promote well-being among surgeons are necessary at the level of surgical residency training in regional areas to recruit surgeons who can work in regional areas.

Although this study revealed the current status of regional disparity in the surgical residency training environment, it has several limitations. First, the response rate of 54% reflects only partial opinions of the surgical residents and is not representative of the total. However, the previous response rate of 56% for the first nationwide survey for surgical residents was comparable to the current study. We believe that it is important to continue conducting surveys and to pay attention to the changes in the results. Second, the RC group in the present study was heterogeneous in that it included cities of various sizes within a population of 1 million. For strict examination of regional disparity, appropriate grouping is necessary based on indicators of rurality such as the rurality index that considers not only the city’s population but also population density, accessibility to medical facilities and so on [25]. Third, revised surgical residency programs were launched in April 2018. Strictly speaking, this system applies to those who obtained their medical license after 2016. In other words, for at least 16% of the UC group and 8% of the RC group residents in this study it was unclear whether they were trained under the previous surgical residency program or the revised one. Therefore, it has been undefined yet what impact the revised surgical residency programs have had on the surgeon shortage in regional areas and the future regional disparity. Continued monitoring surveys to evaluate the surgical residency programs will be necessary. The final limitation is a lack of data on surgical residents who have trained in rural areas that are truly affected by the surgeon shortage and regional disparity. However, under the current surgical residency training programs the completion of surgical residency training solely in rural areas is rare. Nationwide surveys for not only surgical residents but also mid-career or higher surgeons in rural areas who are responsible for the medical facility may help to better understand the current status of community medical care and to address the surgeon shortage with regional disparity.

In conclusion, there was no regional disparity in surgical residency training between urban and regional cities including surgical education and work environment, except for annual income. These findings suggest that surgical residents who trained in regional cities could gain enough surgical experience with a comparable work environment under the current surgical residency training programs compared to those in urban cities. Moreover, this survey also provided hospitals in urban cities with opportunities to improve the training program. These results may lead to encouraging surgical residents to work in regional cities and solve the regional disparity of surgeons in the future. To be cautious, this survey is based on residents’ opinions and not on an objective evaluation of the surgical residency training program. Continuous surveys for surgical residents are necessary to promote the current surgical residency training programs and solve the surgeon shortage with regional disparity.

## Data Availability

All data produced in the present study are available upon reasonable request to the authors

## Acknowledgments

We would like to express our heartfelt gratitude to all the individuals who participated in the survey. We thank Dr. Norihiko Ikeda, former President of the Japan Surgical Society, and other members of the Japan Surgical Society U-40 Working Group who also contributed toward the survey: Yoko Azuma, Tomomi Abe, Keisuke Arai, Sonoko Ishida, Asuka Ishiyama, Momoko Ichihara, Ken Imaizumi, Toshitaka Uomori, Hiroki Uchiyama, Masateru Uchiyama, Daisuke Eriguchi, Kako Ono, Atsuko Kataoka, Yuta Kikuchi, Kensuke Kudou, Takashi Kohmura, Daisuke Koike, Ryosuke Kowatari, Yudai Goto, Yuzuru Sakamoto, Kengo Shibata, Chiaki Suda, Tomoyuki Suda, Shogo Seo, Kosei Takagi, Koji Takada, Shinkichi Takamori, Wataru Takayama, Aya Tanaka, Atsushi Tanikawa, Atsumi Tamura, Takanori Tsujimoto, Koichi Tsuboi, Keita Nakatsutsumi, Mai Nakamura, Akihiro Nagoya, Takeshi Nishimura, Shoko Noguchi, Mariko Fukui, Yuki Fujieda, Atsushi Fushimi, Satoshi Fumimoto, Hideyuki Furumoto, Eisuke Booka, Takahiro Masuda, Tomohei Matsuo, Yutaka Matsubara, Yuichiro Miyake, Yosuke Mukai, Junko Mukohyama, Yuta Murai, Takahisa Yamaguchi, Yoshiko Yamaoka-Fujikawa, Haruhiko Yamazaki, Kyohei Yugawa, Shinichiro Yokoyama, Shiho Yoshida, Kazuki Wakizaka, and Takaaki Watanabe. Further, we would like to thank the Japan Surgical Society Education Committee members, the Japan Surgical Society secretariat staff members, Katrin Beate Ishii-Schrade (Juntendo University) who provided English language editing services.

## Conflict of Interest Statement

T Konishi received grants from Kanzawa Medical Research Foundation, the Japan Kampo Medicines Manufacturers Association, and Pfizer Co. Ltd. outside the submitted work. All other authors have no actual or potential conflict of interest to declare.

## Ethical approval

This research proposal was approved by the research ethics review committee of the Japan Surgical Society (JSS) (JSS2023-1).

## Notes

### Funding Statement

This study did not receive any funding

